# Mental Tasks for Controlling Cursor Movements in a Brain Computer Interface

**DOI:** 10.1101/2023.08.29.23294785

**Authors:** Zahmeeth Sakkaff, Andrew P. Freiburger, Christopher Henry, A. Nanayakkara

## Abstract

Traumatic brain injury and neuro-degenerative diseases leave people dependent and with a low quality of life. Several technologies have been proposed to connect the central nervous system to silicon circuitry and thereby circumvent the damaged nervous tissue that is impairing normal function. These technologies rely upon a language of subtle movements within a patient’s capability to direct computer operations, however, selecting the proper abstraction for even a task as simple as moving a computer cursor can be challenging. Involuntary movements can create noise and false positives for the brain-computer-interface (BCI) receptors, and non-intuitive abstractions can be a barrier for adoption by neurologically damaged patients. We therefore introduce Visualization of Arrow Movements (VAM) as a set of mental tasks for controlling cursor movements in a BCI system. The performance of VAM was evaluated by six untrained subjects via 10-fold cross validation using band power and k-Nearest Neighbor classification methods as well as linear discriminant analysis (LDA) after spatial filtering. The binary classification accuracy in recognizing VAM tasks from each other was between 92% and 100% for four subjects and between 66% and 72% for the other two participants, which suggests that the tasks are most intuitive for even untrained persons. Non-EEG analysis revealed that this performance does not originate from ocular or other facial movements, but from cerebral electrical activity. The high classification accuracy and intuitive abstraction suggest that VAM is a promising abstraction for BCI systems.

## 1 Introduction

Brain Computer Interface (BCI) systems uniquely allow paralyzed patients to communicate with others and thereby radically improve their quality of life [1]. BCI systems control diverse equipment such as prosthetic limbs and wheelchairs with computers that interpret specific mental activity via Electroencephalography (EEG) [2, 3] as computer signals to control machinery [4, 5], as an alternative to more invasive procedures such as those that are being develeoped by Neuralink [6]. BCI relies on the efficiency and accuracy of methods that detect, preprocess, extract a patient’s mental signal [7–10]. These methods can importantly be augmented with Machine Learning algorithms that adaptively sort signal from noise [11]. The selection of imaginary mental task to most effectively transmit as a computer signal [12, 13] – such as multiplying large numbers; tracing a letter; rotating geometric figures; visualizing numbers being counted on a board; visualizing a word; and imaginary limb movements – is therefore imperative for BCI systems, and remains a bottleneck to their adoption. Motor Imagery (MI) [14] – such as imagining hand movements – is the most popular mental task used for controlling cursor movements in BCI and has exhibited high performance and robustness for diverse patients [15], yet it remains susceptible to false positives like other abstractions: e.g. involuntary eye and facial movements [16]. MI abstractions generally, like other mental tasks, do not intuitively relate to the computer operations that they represent, which further hinders their adoption.

We therefore investigated the efficacy of several intuitive MI abstractions – Visualization of Arrow Movements (VAM) – to control a computer cursor. These abstractions further leverage the fact that moving objects stimulate more brain activity than stationary objects, and therefore may be better suited for BCI sensors [17]. Six untrained volunteers employed four VAM that imagined hitting an arrow: from above (to move the cursor down), from below (to move the cursor up), from right (to move the cursor left), and from left (to move the cursor right). The subjects were measured via EEG, similar to other preliminary work on mental tasks [18]. The up and down mental tasks, and the left and right mental tasks, were difficult to distinguished; hence, only the Down Arrow Movement (DAM) and Right Arrow Movement (RAM) in fig. 1 were used for subsequent study, since they are sufficient for screens with continuous borders. Involuntary eye movements were dispelled as a source of overwhelming noise in EEG channels by monitoring eye movements and muscle movements as well as employing spectral analysis and classification techniques. Our results suggest that untrained persons can rapidly master VAM as an intuitive means of controlling a computer cursor, and suggests that this MI has medical utility BCI applications.

**Figure 1:**
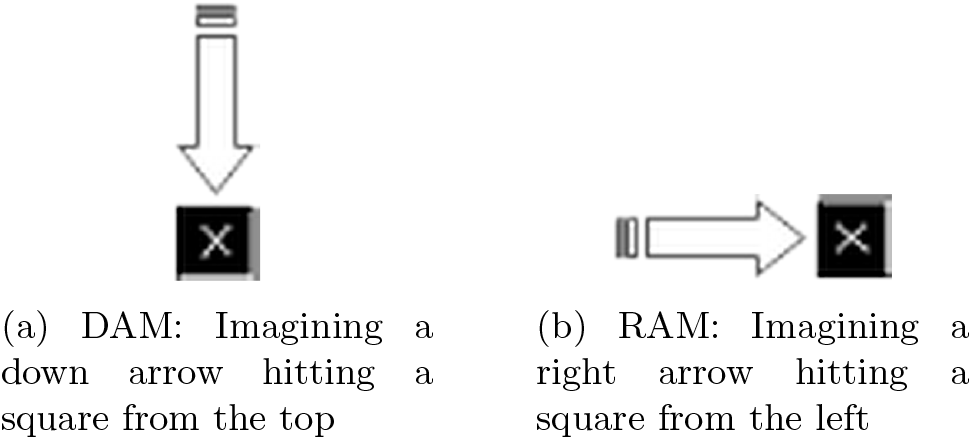
Mental tasks and their associated instructions. This pair of mental tasks was sufficient to localize a cursor anywhere on a computer screen that was programmed with continuous borders.

## 2 Methods

### 2.1 Experimental setup and data acquisition

Six healthy individuals, aged 25-35, were selected to evaluate the performance of VAM. Subjects 2, 3, 4, and 6 (S2, S3, S4, and S6) were male while subjects 1 and 5 (S1 and S5) were female. Subjects S1, S2, S3, S5, and S6 were right-handed and S4 was left-handed. Ethical clearance (2009/EC/36) was obtained for recording of EEG, and volunteer consent was obtained after explaining the experimental procedure and that they will be paid an honorarium for their participation. The Mindset – an 24R amplifier system manufactured by NeuroPulse-Systems LLC, USA – was used to record the EEG signals, which consisted of 24 differential input channels of 90 dB amplifier gain and 60 dB signal-to-noise ratio. ECI Electro-Cap electrode system II manufactured by Electro-Cap International Inc was used to capture EEG signals on the scalp. The EEG electrode placement was based on standard international 10-20 system [19], which includes 20 channels: Fp1, Fp2, F7, F3, Fz, F4, F8, C3, Cz, C4, P3, Pz, P4, T3, T4, T5, T6, O1, O2 and G (ground electrode). Two ear electrodes were also connected to the amplifier as reference electrodes in addition to the electrocap electrodes. The impedance of these electrodes was measured, via an NPS impedance meter (model 1089NP) manufactured by NeroPulse Systems, and maintained for all electrodes *<* 3*K*Ω. A sample rate of 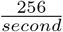 was used.

The role of noise from electrooculography (EOG) and electromyography (EMG) in VAM mental tasks was investigated by examining EOG signals from three different locations surrounding the left eye, since eyes move in unison. Trials where eye blinks or facial movements were observed were discarded, and the affected trial was repeated. The same electrodes for eye movements also monitored involuntary muscles movements such as EOG and EMG artifacts. EEG data was recorded via Mindmeld 24 Data acquisition software, and a program called Alarm was developed to inform each subject about the forthcoming mental task, and when a recording starts and ends, which were signaled via short beeps of different tones. Subjects were primed to visualize the proper VAM before the recording sessions by being shown animated images of DAM and RAM, where a single arrow hits a black cross in fig. 1. Subjects were seated comfortably, with arms and hands relaxed on armrests, during the recording sessions, and looked at a black cross on a white screen ≈ 1.5*m* in front of them to keep their eyes open and minimally moving during the trials. This experimental design in fig. 2 was repeated for 240 trials of each of the three mental task, which resulted in 720 recordings per subject.

**Figure 2:**
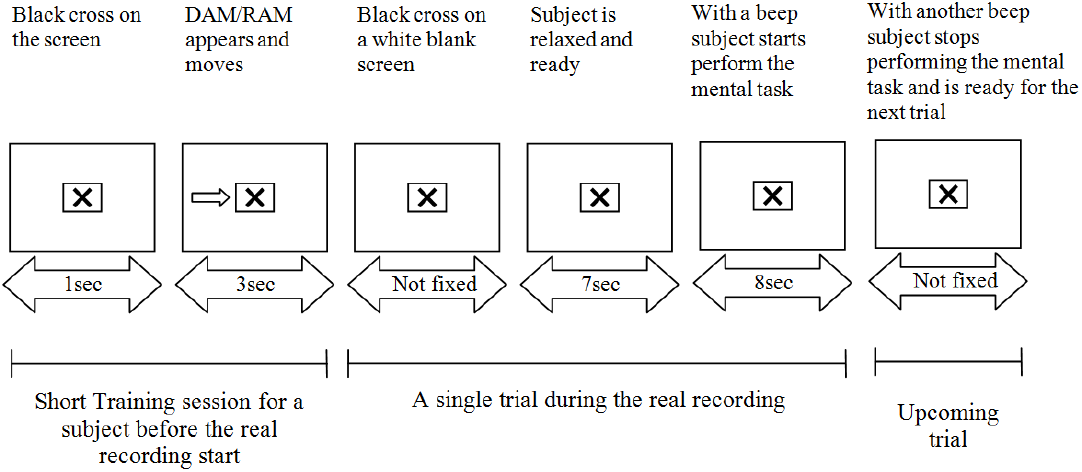
Schematic diagram for the short training session and the real experimental setup showing the time durations for a single trial.

The detectability of EOG signals from eye movements necessitated a separate experiment where subjects performed RAM while moving their eyes side-to-side, performed DAM while moving their eyes up-and-down, and performed both of these VAM mental tasks 1) without moving their eyes, 2) while slightly moving their eyes at a normal speed, and 3) while slightly moving their eyes at a slow speed. EMG was not recorded for facial muscle movements since data was available from literature [16]. The results from these experiments are illustrated in fig. 3.

**Figure 3:**
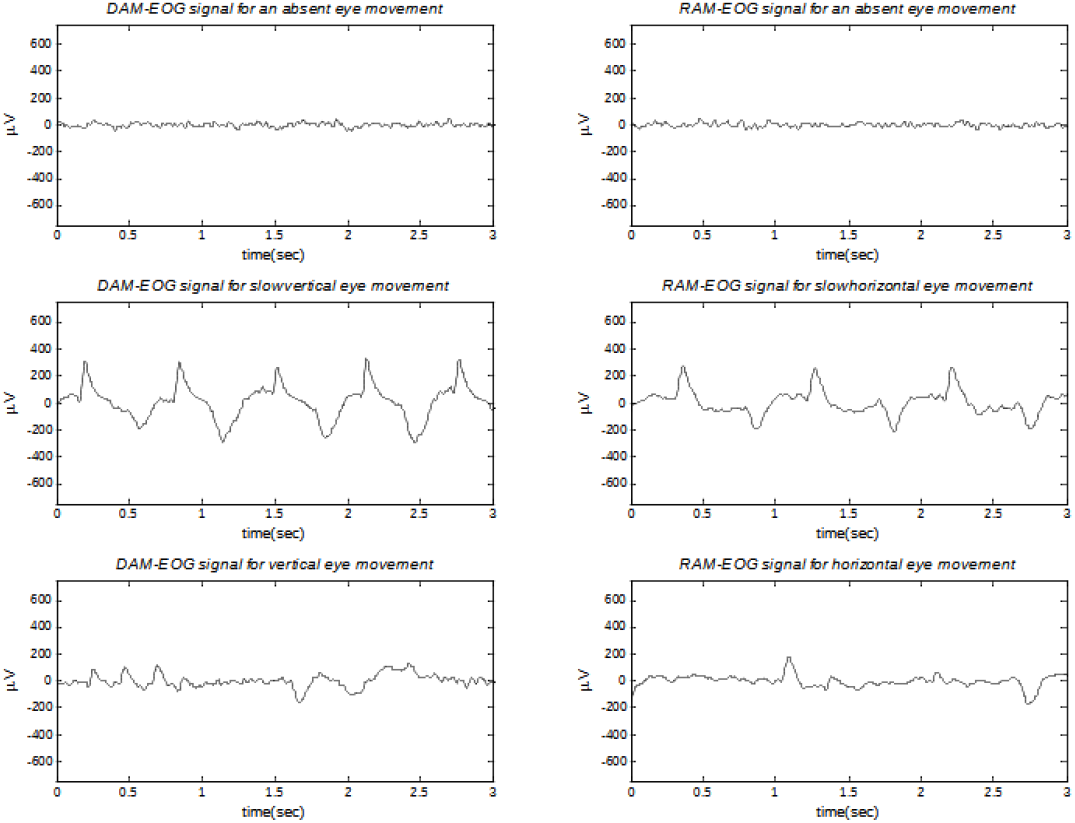
Typical EOG signals recorded for three situations described in the text. Large changes occurred in EOG signals when the subject moves eyes slightly. Even when subject moves eyes slowly, changes in EOG can be easily identified. The above graphs are drawn for the data recorded from subject S1.

### 2.2 Preliminary investigation on signal processing and classification

All calculations were conducted in MATLAB. The EEG signals from each channel were constrained to 1*Hz < freq*_*EEG*_ *<* 48*Hz* with a Butterworth filter that enhances data quality by mitigating noise [20]. This frequency range importantly covers: Delta and Theta ([1, 7] *Hz*), Alpha ([8, 12] *Hz*), Beta ([13, 29] *Hz*) and Gamma ([30, 48] *Hz*) brain frequencies. Test data for feature vector constructions were examined via three methods – Band Power, Down Sampling, and Principal Component Analysis [21]. Band Power, which is based on Fourier Transform, outperformed the other methods, and was utilized for subsequent computations. The cut-off frequencies 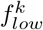 and 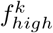 of the *k*^*th*^ band were calculated

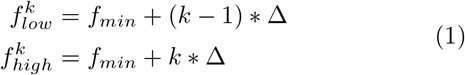

with *f*_*min*_ = 1 *Hz*, Δ as the band width for each partition of the 1*Hz < freq*_*EEG*_ *<* 48*Hz* range, and *f*_*max*_ = 48 *Hz*. Secondly, VAM performance was evaluated by varying between [4, 36] bands for each subject, where 1.3*Hz* corresponded to 36 bands and 12*Hz* corresponded to 4 bands. Common classification methods for evaluating VAM performance were evaluated: linear and non-linear Discriminant Analysis, k-Nearest Neighbor (kNN) classification, and Support vector machines (SVM) with various kernals. Both kNN and SVM exhibited reasonable accuracy with our data, however, some SVM calculations did not converge within a practical number of iterations; hence, kNN was used to assess the performance of VAM in combination with bandpass filtering for preprocessing and Band Power for feature vector construction. The most discriminating information from the VAM mental tasks was furthermore obtained by Common Spatial Patterns and classified with linear discriminant analysis (LDA) [8]. The preliminary classification results reveal that no relative importance could be assigned to any of the spectral bands which are common to all the subjects.

First, the total data set was split: one half was used to identify optimal parameters, such as the most effective channels, bandpower, and classification parameters for each subject; the other half was used to evaluate the performance of VAM. The most effective five EEG channels of each subject are presented in table 1, which represent the fewest channels that still retain classification accuracy. The frequency range [1, 48] *Hz* was used for bandpass filtering and binary classification of VAM, where non-overlapping bands have equal band widths for Band Power in feature vector constructions and 11 nearest neighbors in kNN classification. Performance *P* of each subject with 10-fold cross validation was calculated 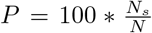 as the percent of the total number of mental tasks *N* that were successfully identified *N*_*s*_.

**Table 1:** The best 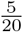 channels for each subject.

| Subjects | Channels |
| --- | --- |
| S1 | *Fp1, Cz, C4, T4, Pz* |
| S2 | *Fp1, Cz, C4, T4, Pz* |
| S3 | *Fp1, F8, C4, T4, Pz* |
| S4 | *Fp1, Cz, F8, T4, Pz* |
| S5 | *Fp1, Cz, C4, T4, Pz* |
| S6 | *Fp2, Cz, C3, T5, P3* |

## 3 Results

### 3.1 Investigation of possible contamination of EEG due to EMG/EOG during VAM

The possible contamination of eye or facial muscles movements in the VAM EEG signals was dispelled through a few steps. First, the EEG and EOG signals were monitored for eye movements or blinks during the recording, with previous EOG recordings in fig. 3 as a reference. This effort was facilitated by eye and facial these movements usually producing large distinguishable signals. Second, spectral analysis of EEG channels, which have classified performance, were used to indicate noise in EOG and EMG, where the presence of such contaminating noise in DAM or RAM is expected to manifest larger peaks throughout the EEG frequency range. The DAM and RAM spectra were therefore compared with the baseline spectra, where no observable difference was observed for any of the EEG channels.

The possibility that subtle eye or facial muscle movements are undetected in EOG or EMG signals and spectra but influence the EEG signals was also considered. The classifications of DAM and RAM using only the EEG channel were compared with those from only the EOG and EMG channels, with separately optimized parameters of bandpass filtering, feature vector construction, and classification for each subject. The best performance for each subject in table 2 ranged between [45%, 69%] in binary classification of DAM and RAM while the EEG channels were much higher, which demonstrates that EEG signals from VAM are not contaminated by eye or facial muscle movements.

**Table 2:**
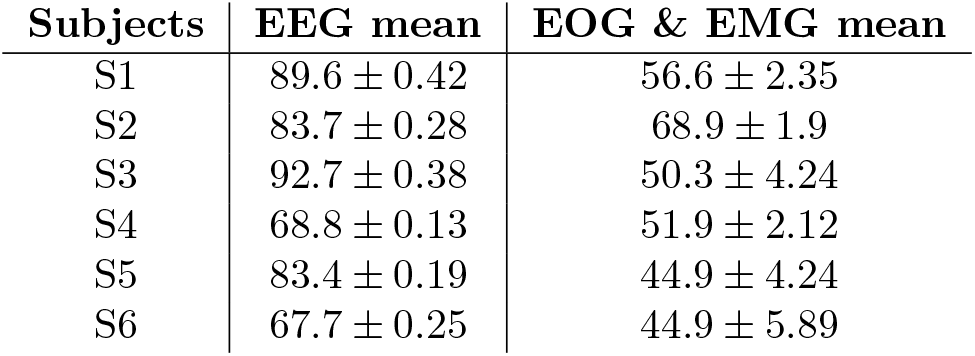
The performance of visualization of arrow movements (DAM vs. RAM) using EGG or EOG + EMG.

| Subjects | EEG mean | EOG & EMG mean |
| --- | --- | --- |
| S1 | 89.6 ± 0.42 | 56.6 ± 2.35 |
| S2 | 83.7 ± 0.28 | 68.9 ± 1.9 |
| S3 | 92.7 ± 0.38 | 50.3 ± 4.24 |
| S4 | 68.8 ± 0.13 | 51.9 ± 2.12 |
| S5 | 83.4 ± 0.19 | 44.9 ± 4.24 |
| S6 | 67.7 ± 0.25 | 44.9 ± 5.89 |

VAM performance from binary classification in table 3 reveals that the classification accuracy of all the subjects except S3 exceeded the baseline versus DAM, in the range [84%, 94%]. and versus RAM, in the range [81%, 93%]. The classification accuracy of RAM versus DAM for the majority of the subjects was similarly between [78%, 87%]; yet, performance for the remaining subjects was low, between [65%, 69%].

**Table 3:** Mean performances of VAM in k-NN based binary classification for channels given in table 2, where BL denotes baseline.

| Subjects | DAM-BL | RAM-BL | DAM-RAM |
| --- | --- | --- | --- |
| S1 | 90.8 ± 0.23 | 86.5 ± 0.19 | 82.6 ± 0.0 |
| S2 | 87.2 ± 0.19 | 88.4 ± 0.46 | 77.8 ± 0.37 |
| S3 | 83.8 ± 0.18 | 85.4 ± 0.37 | 87.2 ± 0.25 |
| S4 | 87.7 ± 0.37 | 82.8 ± 0.35 | 64.6 ± 0.88 |
| S5 | 93.6 ± 0.04 | 92.6 ± 0.18 | 77.7 ± 0.18 |
| S6 | 84.7 ± 0.20 | 80.7 ± 0.28 | 68.7 ± 0.36 |

The head volume effects were reduced via spatial filters to enhance the classification accuracy. Common spatial patterns were compared in table 4 with the support of LDA classifier, similar to other studies. The maximum possible number of filters (m = 10) for each direction were used in the classifier, which significantly improved the classification accuracy from previous studies [22]. The classification accuracy of S1, S2, S3, and S5 were above the baseline between [90%, 100%] for DAM versus [88%, 95%] for RAM. The classification performance of S6 interestingly deteriorated by 11% and 14% for DAM and RAM versus the baseline, respectively, compared to kNN, and the classification accuracies of DAM and RAM for this same subject did not improve from spatial filtering. These anomalous results with S6 are not yet understood; nevertheless, 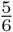 subjects exhibited improvement in the classification accuracy from LDA, which suggests that alternative enhancement methods to spatial filtering may be suitable.

**Table 4:** Classification accuracy for VAM in binary classification using LDA after spatial filtering.

| Subjects | DAM-BL | RAM-BL | DAM-RAM |
| --- | --- | --- | --- |
| S1 | 90.0 | 90.0 | 96.6 |
| S2 | 95.0 | 88.3 | 93.3 |
| S3 | 100.0 | 93.3 | 100.0 |
| S4 | 81.6 | 78.3 | 71.6 |
| S5 | 96.6 | 95.0 | 91.6 |
| S6 | 73.3 | 66.6 | 66.0 |

### 3.2 Potential Physiological Origin of VAM

The priming of subjects with images before recording indicates the possibility of leveraging visual memory and spatial imagery with DAM or RAM, and likely activate the visual cortex, somatosensory cortex, and motor cortex, among other regions of the cerebral cortex. The possible Brodmann areas [23] and their corresponding functions that are activated during VAM are articulated in table 5, which reveals how VAM mental tasks are processed by the brain.

**Table 5:** VAM mental tasks and associated Brodmann areas.

| Brodmann Area | Brain region | Associated activities |
| --- | --- | --- |
| Area 1 | Postcentral Gyrus Primary somatosensory area (SI) | This is the main sensory receptive area for the sense of touch. It also participates in movement organization and performance as well as “mirror neurons” that are vital to understanding anticipation, imitation, and imagery. |
| Areas 5 & 7 | Somatosensory Association Cortex | These areas participate in processing visuo-spatial, activities spatial imagery, mental rotation, similar praxic (motor planning) abilities and imitation of motor learning. |
| Areas 9 & 10 | Dorsolateral / Anterior Prefrontal Cortex | Areas 9/10 have a substantial involvement in memory, particularly memory encoding, memory retrieval, spatial memory and working memory. |
| Areas 17, 18, 19 | Primary (V1), Secondary (V2), Associative (V3) Visual Cortex | In addition to main functions of V1, V2, and V3, these areas also participate in processing of visual patterns, visuo-spatial information, tracking motion, visual attention and visual mental imagery tasks. |
| Area 21 | Middle Temporal Gyrus | It has been reported that this area is associated with function such as observation of motion which is related to VAM. |
| Area 37 | Fusiform Gyrus | This area is participating in functions like visual motion processing and visual fixation |
| Area 40 | Supramarginal Gyrus | Brodmann area 40 is associated in controlling movements guided by visuo-spatial information and complex motor activity such as motor planning plays. |

The EEG channels Fp1, Cz, C4, T4, and Pz in the 10 – 20 system generally exhibited the best classification performance. It is interesting to note that despite a subtle disconnect between electrode activity and brain activity directly below due to volume conduction, the Brodmann areas near these five channels belong to the list of possible Brodmann areas that are associated with VAM.

## 4 Discussion

Brain-computer-interface technologies offer a profound means of improving the lives of paralyzed patients and other persons with debilitating neurological conditions. We investigated several mental tasks that more intuitively relate to moving a computer cursor and require minimal training, which may facilitate adoption by patients. All but one test subjects in our study exhibited *>* 80% accuracy in binary classification of VAM versus the baseline. Potential interference of eye or facial muscles movements in VAM measurement was discredited by monitoring the subjects’ eyes, examining EOG and EMG signals, and classifying DAM and RAM offline using exclusively EOG and EMG signals. Facial movements were monitored through EMG since involuntary muscle movements are only expected on the head. The lack of interference contamination from facial movements is corroborated by spectral analysis of EEG signals, where EOG and EMG noise contamination was not observed for any of the EEG channels in any subject. These observations, in addition to the poor classification results from EOG and EMG signals, indicate that the EEG signals used in classifying of DAM and RAM investigation are not contaminated from EOG and EMG artifacts.

Suitable channels for each subject have to be predetermined at the training stage to practically apply VAM and accommodate subject variability This variability, however, may manifest from insufficient training with the mental tasks. Discrepancy in the accuracy of binary classification of RAM versus DAM may be attributable to the use of k-NN classification after constructing feature vectors with Band Power, since the SVM linear method out-performed kNN for most subjects.

Spatial filters reduced effects from the head-volume conductor and improved local oscillations for motor imagery mental tasks. Spatial filtering also enhanced the classification accuracy significantly for VAM mental tasks: for example, the binary classification accuracy from Band Power and the kNN classifier improved 15% after spatial filtering in four subjects. Several Brodmann areas that may become activated were also identified, although, our limited set of 20 channels was insufficient to localize VAM activity on the scalp. High resolution cortex images from high-density EEG, fMRI or fNIRS during VAM mental tasks would be necessary to localize the neuro-physiology from VAM mental tasks.

The offline study presented here demonstrates the ability of VAM to control both horizontal and vertical cursor movements, and suggests applicability to *in situ* BCI systems. We believe that VAM, as an intuitive mental abstraction of controlling a computer mouse, will be more readily adopted by patients relative to contemporary options and can thereby improve the lifves of paralyzed and neurologically damaged persons. VAM may further be hybridized with other visualizations, such as finger movements, to intuitively simulate clicking mouse buttons and improve signal accuracy, although, this may limit accessibility of the MI for some paralyzed patients.

## Data Availability

All data produced are available online at: https://zenodo.org/record/8264860

## Contributions

**ZS** organized and implemented the study, processed the results, and contributed early writing. **AF** and **CH** contributed writing and organization. **AN** designed the study and secured funding.

### Acknowledgment

This work was funded by the US National Science Foundation through grant MCB-1449014, the National Institutes of Health 11-P20-GM113126-01, hi Sa the NSF EPSCoR EPS-1004094 and the CCF-1161767. Also the grant supported by NIH 11-P20-GM113126-01. The authors would also like to thank KBase developers for their active support throughout the development of this research.

## Conflict of interest

The authors declare no conflict of interest with the presented work.

## Supplementary Information

The raw experimental data and the software that was utilized to process the data are available on Zenodo (10.5281/ zenodo.8264860) as Zip files for others to reproduce our analyses.

## Notes

### Competing Interest Statement

The authors have declared no competing interest.

### Author Declarations

Ethics Review Committee, Faculty of Medicine - University of Peradeniya, Approval number: 2009/EC/36

